# Acute Day Units for mental health crises: A qualitative study of service user and staff views and experiences

**DOI:** 10.1101/2020.11.20.20235374

**Authors:** Nicola Morant, Michael Davidson, Jane Wackett, Danielle Lamb, Vanessa Pinfold, Deb Smith, Sonia Johnson, Brynmor Lloyd-Evans, David P J Osborn

**Affiliations:** Division of Psychiatry, UCL, 149 Tottenham Court Road, London, W1T 7NF, UK; Department of Applied Health Research, UCL, 1-19 Torrington Place, London, WC1E 7HB, UK; McPin Foundation, 7-14 Great Dover St, London SE1 4YR, UK; Camden and Islington NHS Foundation Trust

**Keywords:** mental health, psychiatry, acute care, crisis care, acute day unit, alternative to admission, qualitative, severe mental illness, service users

## Abstract

**Background:** Acute Day Units (ADUs) provide intensive, non-residential, short-term treatment for adults in mental health crisis. They currently exist in approximately 30% of health localities in England, but there is little research into their functioning or effectiveness, and how this form of crisis care is experienced by service users. This qualitative study explores the views and experiences of stakeholders who use and work in ADUs.

**Methods:** We conducted 36 semi-structured interviews with service users, staff and carers at four ADUs in England. Data were analysed using thematic analysis.

**Results:** Both service users and staff provided generally positive accounts of using or working in ADUs. Valued features were structured programmes that provide routine, meaningful group activities, and opportunities for peer contact and emotional, practical and peer support, within a ‘safe’ environment. Aspects of ADU care were often described as enabling personal and social connections that contribute to shifting from crisis to recovery. ADUs were compared favourably to other forms of home- and hospital-based acute care, particularly in providing more therapeutic input and social contact. Some service users and staff thought ADU lengths of stay should be extended slightly, and staff described some ADUs being under-utilised or poorly-understood by referrers in local acute care systems.

**Conclusions:** Multi-site qualitative data suggests that ADUs provide a distinctive and valued contribution to acute care systems, and can avoid known problems associated with other forms of acute care, such as low user satisfaction, stressful ward environments, and little therapeutic input or positive peer contact. Findings suggest there may be grounds for recommending further development and more widespread implementation of ADUs to increase choice within local acute care systems.

## Background

Acute mental health care is provided in the UK in two main forms: acute inpatient psychiatric care, reserved for the most at-risk cases, often with detention under the Mental Health Act; and Crisis Resolution and Home Treatment teams (CRTs) that provide short-term, intensive home treatment to avoid hospital admission or support people at home following an acute admission [1]. There are well-known problems of both these service models, and calls for improvements in UK crisis care, particularly in providing recommended interventions; improving care continuity and outcomes; and reducing reliance on compulsory detention and delays in accessing care [2-6]. These, and other studies [7-9] have highlighted sub-optimal service user experiences of both in-patient and home-based CRT care.

Acute Day Units (ADUs) are a less widely available component of local acute care systems in the UK that, along with crisis houses [10], provide community-based alternatives to hospital admission for those who are not compulsorily detained. ADUs have evolved from acute day hospitals that historically provided non-residential care in some areas the UK, Europe, and the USA to provide an alternative to acute admission, and support into longer term recovery [11, 12]. In their modern incarnation in the UK, ADUs provide a more crisis-focussed and time-limited non-residential service, specifically for adults in mental health crisis who are using or would be considered for acute inpatient psychiatric treatment, or other forms of acute care. They have a similar remit to CRTs within acute care pathways, providing short-term intensive treatment (typically for about a month), and aiming to reduce admissions, or facilitate earlier hospital discharge. ADUs typically provide a range of interventions within their daily programmes, including psychological treatments, practical and peer support, and medication management, with a strong focus on group-based delivery. In the National Health Service (NHS), they are staffed by similar multidisciplinary teams to other acute services including nurses, psychiatrists and support workers, with variable input from other professionals such as psychologists and specialist therapists [13].

ADUs have not been mandated as part of statutory NHS provision but may have the potential to mitigate current problems with both inpatient and CRT treatment, and to enhance choice and service user experience by replacing or augmenting other forms of acute care. While service users and their families often prefer to avoid hospital admission, they commonly report that home-based CRT care provides insufficient input, few therapeutic interventions, and only brief home visits that lack staff continuity [9]. High rates of loneliness are also reported among those leaving CRT care [14].

Research on ADUs is sparse. A UK trial reported symptom improvement and greater satisfaction at discharge for ADU compared to inpatient treatment [15], and a meta-analysis found no differences between day hospitals and inpatient care on readmission rates, life quality, treatment satisfaction and employment, and recommended more and higher quality research [16]. Our more recent mixed-methods programme of research (AD-CARE; Osborn *et al*., in press) explores the provision, effectiveness and acceptability of ADUs in England. An initial mapping exercise in 2016-17 identified 45 ADUs in England, 27 provided within the NHS, and the remainder by voluntary sector organisations, with availability in only 30% of English NHS Trusts (healthcare provider regions) [13]. Our cohort study found no differences in costs, resource use and acute readmissions at 6 month follow-up, greater levels of service satisfaction and well-being, and lower levels of depression in those who used ADUs as all or part of their crisis care, compared to those who only used CRTs [17]. The current paper reports on our exploration of stakeholders’ views and experiences of ADUs using qualitative methods. To our knowledge, there has been no previous research specifically on service users’ experiences of ADUs, or on practitioners’ views of these services. To address this knowledge gap our research aimed to:

i. explore the views of ADU service users, practitioners, and family / informal carers regarding the strengths and weaknesses of ADUs and their component interventions;
ii. explore the views of these stakeholder groups regarding the role of ADUs in the acute care pathway.

## Methods

### Setting

Data was collected in 2017 and 2018 in four NHS-provided ADUs located in urban, suburban and rural areas in the south, south west and midlands areas of England. Detailed descriptions of each site are provided in Appendix 1.

### Patient and Public Involvement (PPI)

Two peer researchers with lived experience of acute mental healthcare and prior experience of qualitative research conducted all interviews, and played primary roles in data analysis. To ensure that relevant experiential perspectives were central to the research process, a Lived Experience Advisory Panel (LEAP) consisting of two additional service users, one family carer, and a PPI expert contributed to all aspects of the project. The LEAP met regularly with other members of the study team and provided input that included: i) contributing to development of interview topic guides; ii) site visits to all ADUs followed by reflective discussion and note-taking to capture impressions of the ‘feel’ of these services; iii) participation in discussions about emerging analytic themes; iv) contributions to developing the focus and orientation of results [18].

### Participant Sampling and Recruitment

Samples of 20 service users, 12 carers and 12 staff members were anticipated to enable access to a diversity of relevant views and stakeholder experiences [19]. We aimed to collect similar amounts of data across the 4 sites. To ensure inclusion of people who broadly reflected the range of ADU service users, we considered age, gender, and ethnicity in constructing the sample. For staff we aimed to include a range of professional disciplines to reflect ADU staff teams as identified in our earlier mapping exercise [13].

Eligibility requirements for service users were: aged 18 or over; having used an ADU for at least one week; understanding of English sufficient to participate in an interview; capacity to provide informed consent; and not posing a high risk to others or themselves. Interested participants were given a study information sheet, and offered a follow-up phone call with a study researcher. Researchers approached ADU managers regarding their own participation, and asked them to identify two more members of the ADU staff team. Carer identification and recruitment was via ADU staff and service users. Written informed consent was provided by all participants.

### Data Collection

Semi-structured interview schedules tailored to each stakeholder groups’ perspective were designed by the research team and the LEAP, and piloted in early interviews (with this data becoming part of the main sample). Some interview questions were broad, to allow discussions guided by participants’ own service experiences; others had a more specific focus on comparisons to other acute services, and ADU activities, culture, and environment. Researchers disclosed their peer status to service user and carer participants in initial contacts to explain the study. Interviews were audio-recorded and transcribed.

### Data Analysis

We analysed data using thematic analysis [20] within NVivo software, adopting a primarily inductive approach. Initially, peer researcher interviewers explored a small sub-sample of data and developed a preliminary coding framework. This was revised and refined through a cyclical process of reading, coding, reflection and discussion with other researchers. Our analytic approach was collaborative throughout, with regular team meetings to discuss analytic processes and emergent themes. These were informed by reflections from members of the LEAP who had visited or had prior experience of ADUs and other acute settings [21]. Emergent themes were organised to relate back to initial research questions in the later stages of analysis.

## Results

### Setting and Interviewee Characteristics

Descriptions of the four services where data were collected are included in Appendix 1. In order to protect sites’ anonymity, they are referred to in this paper as Apple, Cherry, Lime and Peach ADUs.

Interviews were conducted with 21 service users, 7 at Apple ADU, 5 each at Cherry and Lime, and 4 at Peach. Interviewees had attended the ADU for an average of 30 days, 2-5 days per week for an average of 4.5 hours per day. Carer recruitment was problematic, despite attempts to achieve this using multiple methods (via carers groups, ADU staff and service users, and adverts). We recruited three carers, one each from Cherry, Lime and Peach ADUs, all of whom were female and white British. Twelve ADU staff members were interviewed, 3 at each site. This included the manager of each site (usually a psychiatric nurse), nurses, occupational therapists and support workers, who had worked at the ADU for an average of 6.6 years. Further sample characteristics are shown in Tables 1 and 2.

**Table 1.** Demographic and service use characteristics of service user respondents.

|  | **Service users N=21** |  |
| --- | --- | --- |
| Gender | Male | 10 |
|  | Female | 11 |
| Age | 16-24 | 4 |
|  | 25-34 | 5 |
|  | 35-44 | 3 |
|  | 45-54 | 4 |
|  | 55-64 | 4 |
|  | 65+ | 1 |
|  | Mean age: | 41 |
| Ethnicity | White British | 17 |
|  | White Irish | 1 |
|  | White Other | 2 |
|  | Black British | 1 |
|  | Asian British | 0 |
|  | Mixed ethnicity | 0 |
|  | Other ethnic group | 0 |
| Days of ADU attendance at interview: mean (range) | 30 (11-90) |  |
| Days of ADU attendance per week: mean (range) | 4 (2-5) |  |
| Hours of ADU attendance per day: mean (range) | 4.5 (1.5-6) |  |

**Table 2.** Professional and demographic characteristics of practitioner respondents.

|  | **Practitioners N =12** |  |
| --- | --- | --- |
| Role | Manager/nurse | 4 |
|  | Psychiatric nurse | 2 |
|  | Occupational Therapist | 3 |
|  | Support Worker | 3 |
| Gender | Male | 4 |
|  | Female | 8 |
| Age | 16-24 | 0 |
|  | 25-34 | 3 |
|  | 35-44 | 2 |
|  | 45-54 | 4 |
|  | 55-64 | 3 |
|  | Mean age: | 45 |
| Ethnicity | White British | 9 |
|  | White Irish | 2 |
|  | Asian British | 1 |
| Years worked in NHS:<br>mean (range) | 14 (3.5-28) |  |
| Years at current service:<br>mean (range) | 6.6 (0.25-26) |  |

### Qualitative findings

Qualitative findings are structured according to our two broad study aims. First, we explore respondents’ experiences and views of the day-to-day functioning of ADUs. Secondly, views on the role of ADUs within the acute care pathway are presented. For illustrative quotations, the ADU is specified, followed by whether the interviewee was a service user (SU), staff member, or carer. Across both stakeholder groups and sites, ADUs were described positively, especially in comparison to other acute services. With the exception of one service user, all respondents gave broadly positive accounts of using or working in an ADU, in which reported strengths outweighed perceived weaknesses. There was generally common ground of views across stakeholder groups and study sites, although some specific exceptions to this and variations are reported below.

#### 1) Day-to-day functioning and components of ADU care

##### 1.1 The ‘daily-ness’ of ADUs: Routine, purpose, and consistency

The structure and routine of ADU attendance was frequently cited as important, and described by some service users as the most helpful aspect of ADU care. They valued the ‘daily-ness’ of having to be somewhere that encouraged routine, and countered the isolation and lack of structure that sometimes accompany a mental health crisis, and often described this as an important to post-crisis recovery processes. Users also valued the predictable programme of daily ADU activities that were experienced generally as helpful and meaningful.

> Interviewer: *What aspect of this service would you say has been most helpful for you?*
>
> Apple SU2: *I’d probably say the structure and the activities. The coming here each day, having a purpose that’s not too intense, but is engaging enough to kind of stimulate your mind, and having that routine. Because by being at home, you kind of completely fall out of your routine, and you kind of completely fall out of feeling part of society. Whereas coming here, you kind of, you know, it’s almost like going to work*.
>
> *“…so, what it gave me was a structure that I’d been missing for a while. So, routine but also in many ways practical activities that focused on, you know, positive things like recovery and techniques for, you know, dealing with my situation.”* Lime SU5

Some service users contrasted consistent routine and meaningful activities at ADUs to a lack of structure and purpose they experienced both when acutely unwell at home, and in acute in-patient facilities where “*you’re just left to your own devices”* (Lime SU2). Staff also saw structure and routine as an important therapeutic tool. Additionally they highlighted how staff consistency afforded by a weekday, office hours service facilitated continuity and coherent team-working, which is often difficult to achieve in other acute services due to 24 hour shift working:

> *“I think at a fundamental level it sort of provides a kind of structure for people’s day. I think people with … a lot of mental health problems benefit from having a routine of structured activity. That’s aside from all the specific kind of things that we do here.”* Apple staff 2
>
> *“It’s the best team I’ve ever worked with here, because it’s gelled. Because it’s consistent, it’s 9-5 Monday to Friday, so it’s not having to cover those shifts. So you’re on shift with the same staff each day, so the dynamic is constant […]. Whereas when I’ve worked at a crisis team, because it’s shifts, each time you go on it’s a different balance of staff, which creates a bit of a different dynamic, which can be positive or negative in terms of patient care, and in terms of your own stress levels.”* Cherry staff 3

##### 1.2 Group Activities: Meaningful connection and reconnection in post-crisis recovery

Group-based activities were central features of all ADUs studied, providing the principal structure to daily programmes. The diversity of group activities was valued, and they were generally described by users as meaningful to their recovery (sometimes in contrast to the equivalent in acute in-patient settings) in helping to develop peer relationships and provide a focus for their day.

> *“On the ward they’ll do, like, colouring or maybe, like, making something crafty, where here it’s actually helping you, like, with your recovery. I find that they’re giving you, like, recovery sessions and care planning and anxiety management and things like that, and relaxation. And I think it’s all helpful.”* Lime SU2

However, a minority of service users described group participation as challenging, particularly in the early weeks of their attendance when some felt too unwell to participate or benefit from group situations.

The focus of group activities varied between ADUs, shaped in part by staff-team skill sets (see appendix). Staff described designing group activities to interface with local community provision, in order to encourage service users to develop interests and social connections in the community as part of their recovery. Across ADUs, group activities consisted of three broad types:

i. Practical groups focussed on life skills (e.g. cooking, gardening) described by some service users as enabling or confidence-building.
ii. Psychoeducational, therapeutic and self-management groups. Some groups focused on learning about common mental health problems and their management (e.g. anxiety, depression, mindfulness), others had a more general recovery orientation (e.g. ‘moving on’ or care planning groups). These were often described by users as the most valuable groups, enabling them to develop new coping and self-management strategies.
iii. Creative, expressive or well-being focussed groups (e.g. art, music, dance, yoga, pilates, relaxation).

Underlying both service users’ and staff descriptions of group-based activities were concepts of connection and reconnection, with (sometimes implicit) links to recovery processes. Groups were often described as opportunities to connect with others in meaningful ways, through discussion or shared activities, and with aspects of oneself that built confidence or new skills. They were also described as enabling people to reconnect with previous skills or interests they may have lost touch with, or as gateways into social reconnection.

> *“I used to dance quite a lot. I haven’t danced in a long time, but when I came here I did dance therapy and I started dancing again, and obviously that’s beautiful to start dancing again, and I remembered I loved it. And then, like now, in the mornings, when I wake up, I put on some music and I try and dance, and that relaxes me.”* Apple SU2
>
> *“I’ve never done pilates before and it’s really gentle and it’s not very advanced. But it’s a*… *I think it’s a lovely offering that they give, so, yes. And, yes, the other stuff on motivation, planning and, you know, goal setting have also been good to work on. Stuff that I*… *not been there and I’d been avoiding. So, it got me to bring my focus back onto the, you know, pathways to recovery and solutions.”* Lime SU5

##### 1.3 Staff contact: Ad hoc emotional support, signposting, and practical support

All the ADUs studied provided regular meetings with a named key worker and medical input to prescribe, review and monitor medication and conduct physical health checks. Other forums for staff support varied between sites, and included a daily ‘one-stop-shop’ and out-of-hours phone support. Service users generally said little about formal one-to-one staff contact. In comparison, they spoke more about the importance of ad hoc or informal staff contact, particularly in providing emotional support when needed. They valued general staff presence, flexibility and responsiveness.

> *“There was one day in particular where I wasn’t feeling myself and … my mood had dipped. And the staff had noticed this and had a private chat with me about what I was going through at that time. And I was just really appreciative that, A: they had noticed it; but also they were* <u>here</u> *to notice it. And that I had that support and we were able to just kind of tailor things for where I was at that time, and just help me further with my recovery.”* Lime SU3

Staff identified relatively lower caseloads, more contact time compared to other acute services, and staffing consistency without a shift system as enabling them to get to know service users well, develop therapeutic relationships, and provide more meaningful, flexible and personalised support.

> *“I think we manage patients well, and we get to know them so well that you can see if someone’s having a day where they’re not quite themselves. And we can say to them actually are you okay today, you know do you need to chat.”* Cherry staff 3

Both staff and service users identified the importance of support with practical problems. Several staff acknowledged that practical problems and life events or disruptions were often triggered or exacerbated mental health crises, and should be addressed in addition to symptom management. The short-term nature of ADU care meant that signposting to other agencies was often used. Examples of this were described for immediate practical problems (e.g. housing, welfare benefits and legal issues), and longer-term issues relating to recovery, self-management, health and well-being (e.g. alcohol problems, returning to education, exercise). As with group activities, staff recognised the value of building links in the local community to counter social isolation and provide stepping-stones to local non mental health-specific groups or services in the post-crisis period.

> *“One lady has been with us for about four weeks and she really enjoyed knitting on the unit. So, we said oh, why don’t you do a knitting group in the community? And she’s quite isolated – quite socially isolated. So, we were able to find three different knitting/crochet groups that she could do within [the area].”* Peach staff 2

There were also examples at all ADUs of staff providing practical support themselves rather than referring on. This took the form of *“provid[ing] a bit more basic care – mak[ing] sure somebody washes and eats, has clean clothes to put on, that kind of thing”* (Apple staff 2), or help with specific problems (e.g. attending an employment tribunal or a welfare benefits appointment with a service user), and was highly valued by service users.

##### 1.4 Informal peer support: Countering isolation and making connections

Service users and staff across ADUs emphasised the importance of informal peer support to counter to social isolation and loneliness, and enable authentic connections. These occurred through sharing experiences of mental health problems (even if only implicitly acknowledged), and exchanging knowledge of local community resources and self-management strategies, and could inspire hope or help with recovery. Importantly for some, these connections could be free of mental health stigma:

> *“At home I tend to sort of burrow into myself and ruminate, whereas here you get to do the classes, but you also get to speak to the other patients, and that itself is quite therapeutic, just talking and just sharing experiences. Because I tend to find that outside of here, so in terms of the real world, you have to, not put on a façade, but you can’t always be your authentic self, because mental health is stigmatised, or you might have felt stigmatised, and people just don’t understand.”* Apple SU2

Staff emphasised the importance of leaving some parts of the daily programme unstructured to foster peer support and friendships between users, whilst allowing opportunities for staff to provide one-on-one support where needed.

> *“In the morning, we leave the clients for half an hour on their own, to sit and chat to each other without any staff members there. And the clients will tell us that they learned so much from other clients, because they’re saying things like, ‘I thought I was worse than anybody else, and nobody else had these problems. But now I spoke to so-and-so, I’ve realised I can get better.’”* (Cherry staff 1)

##### 1.5 Feeling safe: the reassurance of consistency

Most service users described ‘feeling safe’ at ADUs, especially in comparison to in-patient facilities, often relating this to aspects of service culture and staffing. Feeling safe in relation to other people was linked to features such as a sign-in process (allowing awareness of which staff and service users were present), absence of security wardens, the freedom to come and go, availability and consistency of staff, and being able to trust staff to understand and deal with conflict or challenging behaviour should it arise. Service users appreciated staff checking in with them on bad days to support a sense of safety in relation to their own internal mental states. Overall, consistent and robust staffing and structure appeared to provide reassurance and containment for vulnerable service users.

> *“Some people are going to go off on one, but staff are pretty quick to respond and to kind of know how to, you know, what to say and do. And I feel that they know what to do and say if someone else is kicking off. It rarely happens, but I trust the staff that it will be dealt with and respecting the rest of the people but not taking away the dignity of the person who’s kicked off either. I always felt safe, and I still feel safe now.”* Apple SU1
>
> *“What makes me feel safe? I think it’s the fact that the staff are open and approachable and understanding. It helps create a reassuring environment.”* Lime SU3

Staff spoke about how the voluntary ethos of ADUs contributed to safety and low levels of aggressive or violent incidents. Several staff felt that, compared to in-patient wards, ADUs provided ‘less traumatic’ (Cherry staff 3) or chaotic environments that felt safer for both staff and service users, and were warmer and more caring. Service users being able to take responsibility for their own safety was seen as important.

> *“I think its supportive staff. I think it’s the ability to have an open environment, and a warm and welcoming environment, where people can flourish therapeutically, and not have as much boundaries as they would do on the ward. I think that, in a sense, creates more safety, because they’re still in the community, but they just feel more supported, especially when they’re feel quite vulnerable in their mental health”* Peach staff 2

##### 1.5 Suggested improvements: Longer and more

Responses to a specific interview question on suggested improvements to ADUs reflected the generally positive views expressed across the sample, with five service users unable to think of any improvements, and most suggestions consisting principally of ‘more of the same’ comments. The most commonly suggestion (made by around a third of both service users and staff) was to extend the length of ADU treatment by a few weeks, in order to enable people to stabilise in the post-crisis period in relation to ADU participation or medication changes:

> *“I think it could take at least four weeks to settle in and to get used to the routine and the structure. And once you get settled then it’s time to go. So it’s like, at the point it starts to become helpful is the point where you’ve only got a few weeks left, and you’re being discharged.”* Apple SU5

Some service users reported feeling anxious about ADU discharge, and suggested clearer post-discharge planning, or more post-discharge support via telephone, peer or group-based support. Several respondents linked this to the short length of ADU care, and to difficulties accessing other community mental health services following discharge:

> *“I would like to have something like a staying well group. So when people were discharged there was a touch-base period… most people, not all people, but the majority of people have a good experience here, and being discharged to nothing could be … That’s where I think it all starts to break down because the community services, there’s not enough care resources.”* Lime SU3

Across ADU sites, two thirds of staff, and some service users suggested increased staffing levels in order to offer more and a greater variety of ADU care, particularly group-based activities. Other suggestions included improvements to formalised service user and carer involvement and, from staff, the need for more physical space.

#### 2. ADUs within local acute care contexts

##### 2.1 Comparisons: More input than CRTs; fewer negative experiences than in-patient care; more social than both

Both staff and service users generally thought ADUs were more conducive to recovery than inpatient crisis care. All staff respondents with previous experience of acute in-patient services preferred working in an ADU. Many service users’ negative descriptions of previous in-patient crisis admissions mirrored known short-comings, including staff who were ‘too busy’, ‘patronising’, and difficult to engage with. In comparison, ADUs were described as feeling more relaxed, safer and less chaotic. Similarly, brief home visits often by different staff members provided by CRTs were compared negatively to ADUs.

> “*I think it’s basically a good service, yes, and definitely far more positive than being on the wards and far more positive than using the crisis team.”* Apple SU5
>
> *“With the crisis team it’s maybe not intense enough. You don’t always have the time to spend with people, and they don’t get that peer support either at crisis team. So I think we’re a halfway house between the two* [crisis team and ward], *and I think we almost take the best of both. So keep them at home, but give them more support than just an hour’s visit.”* Cherry staff 3

Positive comparisons with other acute services centred around valued features discussed above, particularly the more social nature of ADUs involving group activities and peer interactions and support; allowing people to remain living at home independently whilst having regular and larger amounts of staff contact; a more consistent staff team; and higher staff to service user ratios.

##### 2.2 Roles: structured post-crisis support and hospital avoidance

The four ADUs studied served multiple acute care roles, providing post-crisis support, replacing in-patient care, complementing home-based CRT care, and providing crisis or relapse prevention support. Provision of ‘step down’ care after a mental health crisis usually followed (and sometimes shortened) a hospital admission, home-based CRT care, or for a minority, crisis house admission. This function was appreciated by service users in helping them transition out of crisis and move towards the next stage of recovery.

> *“I’m just very grateful that there is a place like this and I think I would have been in a bit of a problem if I’d just been discharged by the crisis team and had nothing else to go on to. It’s difficult in everyday life to create a structure, and this place has helped me create a structure so that I can have things to work towards.”* Apple SU7

The role of ADUs in avoiding in-patient admissions, or reducing lengths of hospital stay was similarly important to service users. Staff also cited the value of reducing pressures within local acute care systems, especially within the current UK context of over-stretched mental health in-patient facilities that are often at capacity and unable to respond to need.

> *“If it didn’t exist, I’d be in hospital a lot more, definitely, and I wouldn’t be getting the support I needed in the community. So I’d be inpatient more, and there wouldn’t be the groups run like there is here.”* Lime SU2
>
> [ADU not being there would lead to] *“a larger bed waiting list… more cost, obviously, for the NHS… and then hospital admissions that can be quite lengthy but unnecessary. We’ve definitely prevented a lot of hospital admissions, I think that’s been the main thing. It’s that people have been able to come here instead of hospital.”* Peach staff 2

##### 2.3 Integration: Joint working within acute care; lack of awareness in local mental health systems

The ADUs studied were integrated variously within their local acute care system. Apple ADU had close links with a co-located crisis house, taking a majority of referrals from in-patient services; Cherry took referrals only from the local CRT; Peach was fully integrated with a local CRT, with users often supported simultaneously by both services. Lime accepted referrals from multiple sources including the non-acute sector.

A key area of joint working with other acute services at three ADUs was managing clozapine restarts, which would otherwise necessitate a hospital admission to ensure close medical monitoring. In two ADUs, these service users also attended the full ADU programme. Some staff saw this as a success of ADUs in reducing hospital admissions, although some service users perceived the influx of ‘outsiders’ for clozapine clinic attendance as unsettling or intimidating.

Beyond the acute care sector, staff expressed concerns about low awareness of the ADU among potential referrers from other local mental health services. They cited examples of unnecessary or longer than needed hospital admissions without consideration of ADU care, and lack of staff awareness or understanding of what ADUs can offer. This was voiced across sites, but appeared particularly problematic at Lime, where referrals were not managed by acute care providers:

> *“Sometimes when I look on the bed trackers, I see people on there and I just think, why are they in hospital? We could manage this. We could manage that risk. I know this person. But once they get on to that ward, the ward tends to hold on to them and you can’t get them off again… we really struggle getting people off the ward early.”* Lime staff 1

##### 2.4 Not suitable for all

A small minority of service users described ADU treatment as challenging compared to other treatment options. One person said they would choose in-patient over ADU care, because social anxiety made group activities difficult; another found the freedoms and choices at the ADU initially overwhelming following a long hospital admission. Some staff considered the open and group-based nature of ADUs unsuitable for people were very unwell or with histories of aggression or violence, risk to self, or substance abuse, and described sometimes being selective about who they accepted for ADU care: ‘*making sure we don’t unsettle the status quo of the unit and their therapeutic environment’* (Peach staff 3). A few staff viewed ADUs as less suitable for those with personality disorder diagnoses, stating that they thought the short-term nature of ADUs may exacerbate attachment issues.

## Discussion

Our findings show strong endorsement of ADUs by practitioners and service users. Across sites, respondents talked about broadly similar valued features of ADUs: a daily structured programme; meaningful group activities (including combinations of psychoeducational, therapeutic, symptom-management, recovery-oriented, creative, expressive and life-skill-based groups); greater staff contact time and continuity compared to other forms of crisis care; a sense of ‘safety’; and peer support. Interestingly, other ADU-provided interventions such as medication and physical health reviews did not feature strongly in service users’ accounts. Instead, much of what respondents talked about was underpinned by themes of connection and reconnection, experienced through, for example, basic structure, life-skill focussed groups, help with practical problems, links to resources or groups in the local community, and informal interactions with peers and staff. Many of these were described as tools for recovery from mental health crisis. This suggests that, compared to most acute care services that focus primarily on symptom and risk management, ADUs’ more psychosocial and holistic package of crisis care is particularly valued. These findings may help account for positive outcomes in our related quantitative study that reports higher service satisfaction and well-being, and less depression following crisis care at the same four ADUs compared to with CRTs [17].

A key strength of ADUs compared to home-based crisis care is the provision of regular social contact outside the home. For those living alone or without employment or meaningful support from friends or family, these issues can be particularly significant during a mental health crisis. Loneliness and social isolation are common amongst those using crisis services, and are associated with poorer outcomes [22]. ADUs’ facilitation of (re)connections with local community resources beyond the mental health system may enable users to build social and personal capital, and develop broader support and self-management strategies.

Two commonly reported problems with home-based CRT treatment are lack of staff continuity and limited contact time (different staff members making short home visits focussed on risk management) [9]. For hospital-based crisis care, users often report finding the environment chaotic and unsafe, and a lack of meaningful therapeutic activities and engagement with staff [23-25]. Our analysis suggests that ADUs avoid these problems, achieve a more personalised style of care, and are often compared favourably to other forms of acute care. ADUs are neither as constraining as in-patient care, nor as potentially isolating as home-based care. People in, nearing or recovering from mental health crisis can retain the independence of living at home, whilst also receiving higher levels of therapeutic input, and support from staff and peers as part of a structured daily timetable. For practitioners, standard opening hours (rather than 24/7) without a shift system are valued for enabling team cohesion, more contact time, and opportunities to develop therapeutic relationships. However, both groups comment that relatively short lengths of stay (averaging about a month) may limit the benefits of ADU, especially when there is little community-based therapeutic input available post-discharge. Additionally, it should be acknowledged that ADUs are not suitable for people who are compulsorily detained or for whom risk is considered high.

### Strengths and limitations

This is the first study to explore in depth the strengths, weaknesses and acute care role of ADUs from the perspectives of those who use and work in them. It combines stakeholder perspectives across four sites, and benefitted from strong public and patient involvement throughout. Data were collected by peer researchers with lived experience of acute services who played central roles in data analysis and reporting. Our LEAP reviewed study materials, and their impressions from visits to ADUs, and reviews of emerging themes helped shape our findings. This strengthens the study’s understanding of the concerns and perspectives of users and carers, although none of the people working on this study had direct experience of ADU use.

Our recruitment processes via ADUs may have inadvertently produced samples of service users and practitioners with particularly positive views of ADUs. Thus, findings may not fully capture the views of people with less positive experiences of these services. Recent research suggests that views about acute mental health care expressed on social media are generally more negative than those obtained via interviews [9], and recommends using analysis of social media forums to access the views of people who do not engage with statutory services [26].

We were unable to recruit a meaningful sample of family carers, despite using multiple recruitment channels. We interpret this as partly indicative of ADU users often being socially isolated with little regular informal support from family or friends, and of generally low levels of carer engagement at ADUs (only one site provided formal family carer support via monthly groups that were reportedly poorly attended). Carer interviews were short and respondents simply didn’t appear to know much about the ADU their relative attended. However, difficulties in carer recruitment to mental health research are common, and not unique to this study. Additionally, we recruited very few people from black and minority ethnic (BAME) groups. Although this may in part reflect the demographic make-up of catchment areas where we recruited, it is important given that BAME groups face significant inequalities in help-seeking for mental illness [27]. We were not able to purposively select the services where data were collected, and relied instead on those willing to participate, although no significant service design or outcome differences between these and other NHS-provided ADUs were identified in our mapping exercise [13].

### Clinical and research implications

This study and our related cohort study [17] suggest that ADUs can play an important and distinctive role in acute care systems, providing a care model that is popular among those who use and work in them, and is compatible with national service improvement policies [28]. ADUs currently exist in only around 30% of NHS healthcare regions in England and their non-mandated status renders their existence patchy and fragile, with closures and service reconfigurations common [13]. To illustrate, one ADU that was originally part of this study closed abruptly before data collection could commence, and another included in the study closed soon after. Such processes impede the ability of ADUs to become established and to contribute consistently to local acute care networks. Further research including economic evaluations are warranted to potentially strengthen their evidence-base. In particular, system-level evaluations are needed to explore service configurations within local acute care networks that offer choice and good outcomes for the full range of people experiencing mental health crises. Given well-documented pressures on mental health acute wards (CQC, 2017) and high readmission rates [29], investment is needed in a range of non-hospital-based acute care options. But greater understanding is needed of potential knock-on or unintended consequences on the staffing and population of acute hospital wards that remain the only option for those with very high levels of risk or disturbance, or who are compulsorily detained [30]. Such research could inform national guidance and potentially provide a more secure status for ADUs.

However, wider implementation or mandated status may bring other challenges. ADUs may not be suitable for all who are in mental health crisis. Their small scale and ability to offer personalised care that are valued by service users may be compromised. This study goes some way to identifying the critical ingredients of ADU care from the perspectives of service users and practitioners. This could inform an evidence-based model of best practice for ADUs, and a fidelity measure to guide and assess ADU implementation [31]. Integration with other services, service awareness among other local healthcare providers, and an ability to combine role clarity and adaptability have been identified as facilitators of successful implementation and sustainability of community-based acute care services [32]. Between the four ADUs in this study there were variations in roles as alternatives or additions to CRT care or hospital admissions, with service users particularly valuing their role in post-crisis support, often with concurrent CRT care. A previous review [16] estimated that 1 in 5 hospital admissions might be avoided and effectively managed by an ADU. However, in our study practitioners expressed concerns about low awareness of ADUs among other local mental healthcare providers.

Some service users and practitioners in our study felt that length of stay at ADUs was too short, and that this reduced their benefit for some users who found engagement difficult in the early stages of attendance. This study focusses on NHS-provided ADUs, but similar services are provided by the voluntary sector or by NHS-voluntary sector partnerships [13]. These typically provide more socially oriented and relational styles of care with a less medical focus, and are valued by users for being more responsive and flexible than NHS crisis services [33]. A significant difference is their support over longer periods, spanning more of a mental health crisis trajectory from emergence through to later stages of post-crisis recovery. Post-crisis is an important transition period in which people typically remain vulnerable, but can benefit from peer support in developing self-management to aid longer-term recovery and reduce relapse risks [34]. Increased sharing of expertise, and joint working between statutory and voluntary sector providers, with associated evaluation research, could enhance choice, help develop innovative forms of crisis management, and complement the medical and risk-focused approach of most statutory acute care services.

## Conclusions

ADUs are relatively uncommon and under-researched. This qualitative study suggests they provide a form of crisis care that is valued by service users and ADU practitioners, and avoids many of the shortcomings of both home-based and in-patient acute care. Their daily structure and a broad range of therapeutic provision oriented by a psychosocial and holistic view of crisis management is valued in enabling personal and social connections and post-crisis recovery. Findings suggest there may be grounds for recommending more widespread implementation to increase choice and provide a distinctive contribution to local acute care service networks.

## Data Availability

The anonymised dataset generated and analysed in this study are available from the corresponding author on reasonable request.

## List of abbreviations

ADU: Acute Day Unit
CRT: Crisis Resolution and Home Treatment team
LEAP: Lived Experience Advisory Panel
NHS: National Health Service
PPI: Patient and Public Involvement
SU: Service User

## Declarations

### Ethics approval and consent to participate

All procedures contributing to this work complied with the ethical standards of the English National Health Service. Ethics approvals were gained from London Bloomsbury Research Ethics Committee (ref: 16/LO/2160). Informed written consent was obtained from all individual participants included in the study.

### Consent for publication

Not applicable.

### Competing interests

The authors declare that they have no competing interests.

### Funding

This project was funded by the National Institute for Health Research Health Services and Delivery Research Programme (project number 15/24/17). The views and opinions expressed herein are those of the authors and do not necessarily reflect those of the HS&DR Programme, NIHR, NHS or the Department of Health and Social Care. NM, SJ, BLE and DO are supported by the National Institute for Health Research (NIHR) Biomedical Research Centre (BRC) at University College London Hospitals (UCLH). DO is also supported by the National Institute for Health Research ARC North Thames.

### Authors’ contributions

MD and JW collected data for this study. Data were analysed by MD, NM and JW. VP and DS were part of the LEAP who contributed to data analysis and other aspects of the study as detailed in the text. DL managed the study. DO was the Principle Investigator for the research. DO, BLE, SJ, NM and VP conceived and designed the study, and obtained funding. NM drafted the manuscript and all authors contributed to and approved the final manuscript.

## Acknowledgements

We thank members of our LEAP, Hameed Khan, Terry Harper and Deb Smith who provided valuable contributions to this work. We are grateful to all the individuals (service users, ADU staff and family carers) who participated in this research.

## Appendix 1: ADU site descriptions

Site descriptions are based on combined information drawn from our previous national survey of ADUs in England [13], field notes and researcher and LEAP member impressions from site visits, and other material such as operational policy documents. They are designed to provide contextual background about each service to inform readers’ understanding of findings presented in the Results. In order to protect service anonymity, pseudonyms for each service are used.

### ‘Apple’ ADU

Apple ADU has a caseload of about 25 and serves an inner-city area covering both highly deprived and wealthy areas. It is located in a freestanding building on a larger psychiatric hospital site. There is a crisis house on the same site, users of which can attend the ADU. Apple service users are more likely than users of other ADUs nationally to be from minority ethnic backgrounds, have problems with practical issues such as housing and debt, and to have diagnoses of severe and enduring psychosis. Most referrals are from acute psychiatric in-patient services. An average length of stay of 50 days was reported.

Apple ADU offered a diverse and strongly arts-based programme, including dance therapy, drama, acupuncture, aromatherapy and music-based groups. It was the only site to employ part-time arts, music and movement therapists. At the time of research, Apple ADU had operated for 15 years, making it the longest-running study site. It had evolved from a day hospital providing longer-term treatment since the early 1990s. Some staff members were long-serving, and had seen a transition from a predominantly psychodynamic approach to a more recovery-focused, short-term one. During site visits, researchers noted a sense of resistance to change among some staff members. There was a threat of closure, and loss of some ADU space during the study period which impacted negatively on staff morale. Compared to other study ADUs, researchers and LEAP members noted that Apple seemed consistently poorly attended, with sometimes only a handful of service users present. Survey data confirmed high (often over 50%) DNA rates.

### ‘Cherry’ ADU

Cherry ADU is situated in a commuter town close to a medium-sized city. It serves a largely rural and semi-rural catchment area that is relatively deprived compared UK norms and the other rural area ADU in the study, Lime. The service is located on a psychiatric hospital site within the main hospital building, which is grand, early-20^th^-century, and red-brick. Its interior though contrasts with this: The ADU relocated to these newly refurbished premises shortly after the study started. The unit was bright, clean and freshly painted, with service user art displayed throughout, and modern features such as a ‘relaxation room’ with large leather massage chairs and a sound system.

Cherry ADU had been in operation for seven years when data were collected. Referrals came primarily from local crisis resolution and home treatment teams (CRT). People with dementia or primary substance misuse diagnoses were not accepted, and a 30 day average length of stay was reported. The daily programme had a strongly creative focus, providing groups that were more activity-based than explicitly therapeutic or psychoeducational. The unit includes a fully-equipped medical clinic where new service users are given a full physical examination, and a weekly clozapine titration clinic is held. As users of other services attend the clozapine clinic with no additional staffing, the ADU is busier on clinic days and staff are drawn away from other ADU activities to manage this. Compared to other study sites and ADUs in England, Cherry has a smaller annual budget. Researchers became aware during site visits of significant budgetary constraints and related staff concerns. They also noted a highly dedicated staff team, some of whom raised funds for the ADU in their spare time.

### ‘Lime’ ADU

Lime is located in a commuter town, and like Cherry ADU, serves a largely rural catchment including small and large towns; unlike Cherry ADU, the surrounding area is relatively affluent. The ADU is situated on a larger community healthcare site in a quiet residential location. It occupies a modern purpose-built building that researchers described as spacious-feeling, well-presented and functional.

Lime ADU had been in operation for six years when data were collected. Its programme focussed primarily on psychoeducational and psychological groups. Referrals were accepted from multiple sources including the non-acute mental health sector. Lengths of stay averaged 30 days. Compared to other study ADUs, Lime reported the lowest proportion of service users experiencing psychosis, and serves more people with diagnoses of depression or anxiety than other study ADUs. People with dementia diagnoses were not accepted. Reflecting this client group, Lime was the only study ADU that did not provide a clozapine clinic, although it did include a clinic room for physical health checks. It is relatively well-funded and well-staffed, and was the only service to report in our survey that staffing felt sufficient.

### ‘Peach’ ADU

Peach ADU is located near the centre of a large and relatively affluent city. The unit is based on an expansive site, in a modern building also containing mental health in-patient facilities. As we started study recruitment Peach ADU opened for a 6-month pilot, intended to establish the feasibility of developing several larger ADUs across the local healthcare region. It was consequently much smaller than other ADUs in the study, both in terms of caseload (maximum of 10 compared to ∼30 at the other sites) and physical size (comprising only three communal areas and a garden, without dedicated space for one-to-one work).

Peach ADU is jointly run, and fully integrated with the local CRT; accepts referrals only from the crisis team or inpatient wards, rather than from community teams; and all people using the ADU are also on the CRT caseload, often being treated simultaneously by both teams (for example, the CRT might do an evening home visit to deliver medication after the service user had spent the day at the ADU). Peach’s group programme consisted principally of psychoeducational and psychological groups focussed on coping strategies and symptom management. It was the only study ADU to include formal peer support worker roles. Two peer support workers attended the unit for half a day a week each in a voluntary capacity. Peach ADU closed soon after data collection for this study.

## Notes

### Competing Interest Statement

The authors have declared no competing interest.

